# JAK signaling was involved in the pathogenesis of Polymyalgia Rheumatica

**DOI:** 10.1101/2022.03.10.22272242

**Authors:** Fan Yang, Xinlei Ma, Bei Xu, Yiduo Sun, Mengdi Jiang, Chunyun Ren, Chuanying Sun, Junyu Liang, Heng Cao, Danyi Xu, Lihuan Yue, Liqin Xu, Jin Lin, Weiqian Chen

## Abstract

**Objectives:** Polymyalgia Rheumatica (PMR) is a common inflammatory disease in elderly persons whose pathogenesis is unclear. We aimed to explore the pathogenetic features of PMR and find a new therapeutic strategy.

**Methods:** We included 11 patients with PMR and 20 age-matched and sex-matched healthy controls (HC) in this study. The disease features were described. The gene expression profiles were analyzed in peripheral blood mononuclear cells (PBMCs) by RNA sequencing and were confirmed by RT-PCR. We also tested gene expression profiles in five patients with PMR after tofacitinib therapy.

**Results:** Patients with PMR experienced pain with high disease activity scores. The gene expression of PBMCs in patients with PMR differed from that in HC by RNA sequencing. GO and KEGG analysis demonstrated that inflammatory response and cytokine-cytokine receptor interaction were the most remarkable pathways. There were markedly expanded IL6R, IL1B, IL1R1, JAK2, TLR2, TLR4, TLR8, CCR1, CR1, S100A8, S100A12, and IL17RA expressions. Those genes may trigger the JAK signaling. Furthermore, tofacitinib, a pan JAK inhibitor, effectively treated five patients with PMR, leading to clinical remission and a significant decrease in inflammatory genes.

**Conclusions:** Many inflammatory genes associated with JAK signaling were increased in patients with PMR, suggesting an important role of JAK signaling in PMR disease development. JAK inhibitors may effectively treat PMR.

**Key messages:**

1. Patients with PMR had significant inflammatory genes expression. JAK signaling may be highly activated.
2. Tofacitinib may treat PMR with clinical remission and a significant decrease in inflammatory genes.

## Introduction

Polymyalgia Rheumatica (PMR) is an inflammatory disease in elderly patients (>50 years old) characterized by pain and early morning stiffness in the shoulder, neck, and pelvic girdle[1, 2]. Blood tests show higher serum levels of inflammatory markers such as c-reactive protein (CRP) and erythrocyte sedimentation rate (ESR) in these patients; however, negative results of rheumatoid factor (RF), and anti-citrullinated protein/peptide antibody (ACPA), anti-nuclear antibodies (ANA), and human leukocyte antigen B27 (HLA-B27). Sometimes, it is difficult to distinguish patients with PMR from those with Rheumatoid arthritis (RA) who are negative for RF or ACPA. However, patients with PMR usually have hallmark features with a dramatic response to glucocorticoids.

PMR is a multi-gene susceptibility disease, and many alleles may be involved in the pathogenesis of this disease[3]. Interleukin 6 (IL6) polymorphisms are associated with PMR risk or severity[4]. Aging is a crucial element as PMR occurs in elderly patients. Aging may increase the susceptibility to infectious agents, immune responses, or the release of cytokines[5]. However, its pathogenesis has not been fully elucidated. Further pathogenetic research and the discovery of new targets can provide more ideas for the treatment of PMR.

## Methods

### Subjects

This cohort study included PMR patients who fulfilled the 1982 Chuang criteria[6] between September 2020 and September 2021. They did not receive any glucocorticoids or biological agents during the three-month period that preceded their inclusion in the study. There were twenty age-matched and sex-matched healthy controls (HC) without connective tissue disorders, neoplasms, or current infections. All patients gave their written informed consent. The study was performed according to the recommendations of the Declaration of Helsinki and was approved by the Medical Ethical Committee of the First Affiliated Hospital, Zhejiang University (IIT20200070C-R1).

### Clinical features

Patient information on demographic data, clinical features, serological profiles, and medications was obtained from medical records. Current PMR disease activity was measured using the previous score sheet[7].

### RNA sequence analysis and Semiquantitative Real-Time PCR

Peripheral blood mononuclear cells (PBMCs) were isolated from patients with PMR and HC. Total RNA and RNA-seq libraries were prepared. Sequencing was performed on an Illumina HiSeq1500. Some genes were tested by semiquantitative Real-Time PCR. The primers for the target gene were described as below.

IL6R Forward: 5’-CCCCTCAGCAATGTTGTTTGT-3’

Reverse: 5’-CTCCGGGACTGCTAACTGG-3’

IL1B Forward: 5’-ATGATGGCTTATTACAGTGGCAA-3’

Reverse: 5’-GTCGGAGATTCGTAGCTGGA-3’

Interleukin 1 receptor type 1 (IL1R1)

Forward: 5’-ATGAAATTGATGTTCGTCCCTGT-3’

Reverse: 5’-ACCACGCAATAGTAATGTCCTG-3’

Janus kinase (JAK) 2

Forward: 5’-TCTGGGGAGTATGTTGCAGAA-3’

Reverse: 5’-AGACATGGTTGGGTGGATACC-3’

Toll Like Receptor 2 (TLR2)

Forward: 5’-ATCCTCCAATCAGGCTTCTCT-3’

Reverse: 5’-GGACAGGTCAAGGCTTTTTACA-3’

TLR4 Forward: 5’-AGACCTGTCCCTGAACCCTAT-3’

Reverse: 5’-CGATGGACTTCTAAACCAGCCA-3’

TLR8 Forward: 5’-ATGTTCCTTCAGTCGTCAATGC-3’

Reverse: 5’-TTGCTGCACTCTGCAATAACT-3’

Interleukin 17 Receptor A (IL17RA)

Forward: 5’-GCTTCACCCTGTGGAACGAAT-3’

Reverse: 5’-TATGTGGTGCATGTGCTCAAA-3’

Complement receptor 1 (CR1)

Forward: 5’-AGAGGGACGAGCTTCGACC-3’

Reverse: 5’-TCAGGACGGCATTCGTACTTT-3’

C-C motif chemokine receptor 1 (CCR1)

Forward: 5’-GACTATGACACGACCACAGAGT-3’

Reverse: 5’-CCAACCAGGCCAATGACAAATA-3’

S100 calcium-binding protein A8 (S100A8)

Forward: 5’-ATGCCGTCTACAGGGATGAC-3’

Reverse: 5’-ACTGAGGACACTCGGTCTCTA-3’

S100A12 Forward: 5’-AGCATCTGGAGGGAATTGTCA-3’

Reverse: 5’-GCAATGGCTACCAGGGATATGAA-3’

GAPDH Forward: 5’-GGAGCGAGATCCCTCCAAAAT-3’

Reverse: 5’-GGCTGTTGTCATACTTCTCATGG-3’,

GAPDH was considered as a normalization control. The data were examined using the 2^-ΔΔCT^ method and results were expressed as a fold increase. Each sample was tested in triplicate, and tests were repeated three times.

### Statistical analysis

Results are expressed as median and standard deviation (SD). Comparisons between two groups were performed using the nonparametric Mann-Whitney test. *P*-values < 0.05 were considered statistically significant. All statistical analyses were performed using SPSS software, version 18.0.

## Results

### The patients with PMR were in a state of high inflammation

The baseline characteristics of the 11 PMR patients and 20 healthy controls are presented in the supplementary material. There were no significant differences in age and sex between the PMR group and HC. In the PMR group, patients who experienced pain in the shoulder, neck, and pelvic girdle had a mean VS score of 4.9 ± 2.2. They had higher serum levels of CRP (3.9 ± 3.1 mg/dl, normal range 0–0.08) and ESR (67.1 ± 24.9 mm/L, normal range 0–20). Disease activity scores[7] ranged from 9.33 to 27.15 with a mean value of 16.3 ± 7.3 (Table 1), which suggests medium or high disease activity in these patients who were categorized as active PMR (PMR-A). They were all negative for RF, ACPA, ANA, and HLA-B27. They represented no remarkable wrist, metacarpophalangeal, proximal interphalangeal, knee, and digital joint swelling.

**Table 1.** Demographic and clinical parameters of PMR and healthy controls on study entry

|  | PMR-A<br>(*n* = 11) | Healthy<br>controls<br>(*n* =20) | *P* value |
| --- | --- | --- | --- |
| Age, mean ± SD years | 68.0±8.3 | 63.7±9.8 | 0.142 |
| Female/male | 10/1 | 17/3 | 0.67 |
| Disease duration,<br>mean ± SD months | 10.4±14.8 | NA | |
| Pain, VS score 0-10,<br>mean ± SD | 4.9±2.2 | NA | |
| Physician global<br>assessment, VS score<br>0-10, mean ± SD | 4.7±2.9 | NA | |
| Morning stiffness,<br>min, mean ± SD | 17.3±21.8 | NA | |
| EUL, mean ± SD | 1.0±0.8 | 0 | |
| CRP, mean±SD mg/dl | 3.9±3.1 | 0.2±0.2 | <0.0001 |
| ESR, mean±SD mm/h | 67.1 ±24.9 | 10.5±5.2 | <0.0001 |
| Disease activity<br>scores, mean±SD | 16.3±7.3 | NA | |

these PMR patients with high disease activity: PMR-A, NA: not applicable, VS: visual analogue scale, EUL: ability to elevate the upper limbs; C-reactive protein (CRP), normal range 0–0.08 mg/dl; Erythrocyte sedimentation rate (ESR), normal range 0–20mm/h.

### Patients with PMR had significant inflammatory gene expression profiles

The principal component analysis (PCA) and heatmap showed that gene expressions of PBMCs in patients with PMR-A differed from those of HC (Fig. 1A, B). GO analysis demonstrated that the inflammatory response (FDR 6.74 × 10^−15^, included 58 significantly altered genes), plasma membrane (7.30 × 10^−12^, included 254 significantly altered genes), integral component of the plasma membrane (5.23 × 10^−11^, included 115 significantly altered genes), extracellular space (2.76 × 10^−8^, included 103 significantly altered genes), extracellular region (4.36 × 10^−8^, included 116 significantly altered genes), response to lipopolysaccharide (2.42 × 10^−6^, included 27 significantly altered genes), integral component of membrane (1.36 × 10^−5^, included 270 significantly altered genes), immune response (1.21×10^−4^, included 42 significantly altered genes), cell surface (2.04 × 10^−4^), innate immune response (4.29 × 10^−4^), response to glucocorticoid (7.43 × 10^−4^), IL-1β secretion (1.67 × 10^−3^) were 12 significant predominant pathways in PMR (Fig. 1C). The cytokine-mediated signaling pathway, the positive regulation of IL-6 production, and the MyD88-dependent toll-like receptor (TLR) signaling pathway were also significantly altered. KEGG analysis revealed that the cytokine-cytokine receptor interaction was the most remarkable pathway (FDR 2.85 × 10^−5^) (Fig. 1D). GO and KEGG results demonstrated patients with PMR had significant inflammatory gene expression profiles.

**FIG. 1.**
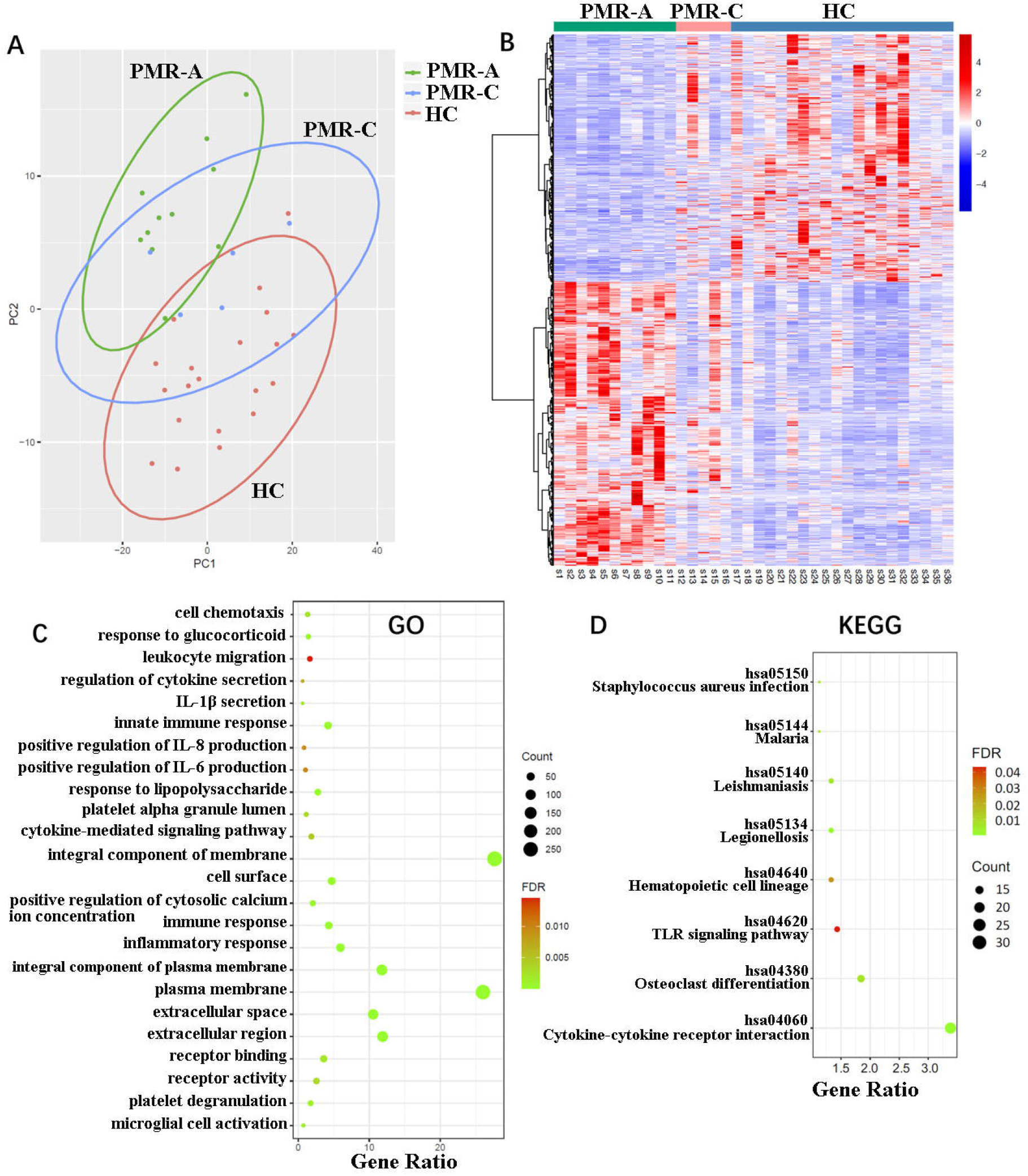
The patients with PMR had a significant inflammatory gene expression profile. **(A and B)** Principal component analysis (PCA) and heatmap showed that gene expression of PBMCs was different among patients with active PMR (PMR-A, n=11), with PMR in remission (PMR-C, n=5), and Healthy controls (HC, n=20) by RNAseq analysis. **(C)** GO analysis demonstrated that inflammatory response (FDR 6.74×10^−15^, 58 significantly altered genes) and other 11 predominate pathways (PMR-A *vs*. HC). **(D)** KEGG analysis showed cytokine-cytokine receptor interaction was the most remarkable pathway (FDR 2.85×10^−5^, 33 significantly altered genes) (PMR-A *vs*. HC).

### Abundant inflammatory genes that may activate Janus tyrosine kinase (JAK) signaling

Volcano results demonstrated that many genes expression were significantly increased; for example, TLR8 (*p* = 1.95 × 10^−20^), IL17RA (*p* = 3.27 × 10^−17^), IL6R (*p* = 6.71 × 10^−16^), CR1 (*p* = 1.14 × 10^−14^), TLR2 (*p* = 2.99 × 10^−14^), TLR4 (*p* = 8.48 × 10^−14^), CCR1 (*p* = 7.75 × 10^−13^), JAK2 (*p* = 6.57 × 10^−6^), IL1R1 (*p* = 1.72 × 10^−5^), IL1B (*p* = 2.87 × 10^−5^), S100A8 (*p* = 2.14 × 10^−3^), and S100A12 (*p* = 2.76 × 10^−3^) (Fig. 2A).

**FIG. 2.**
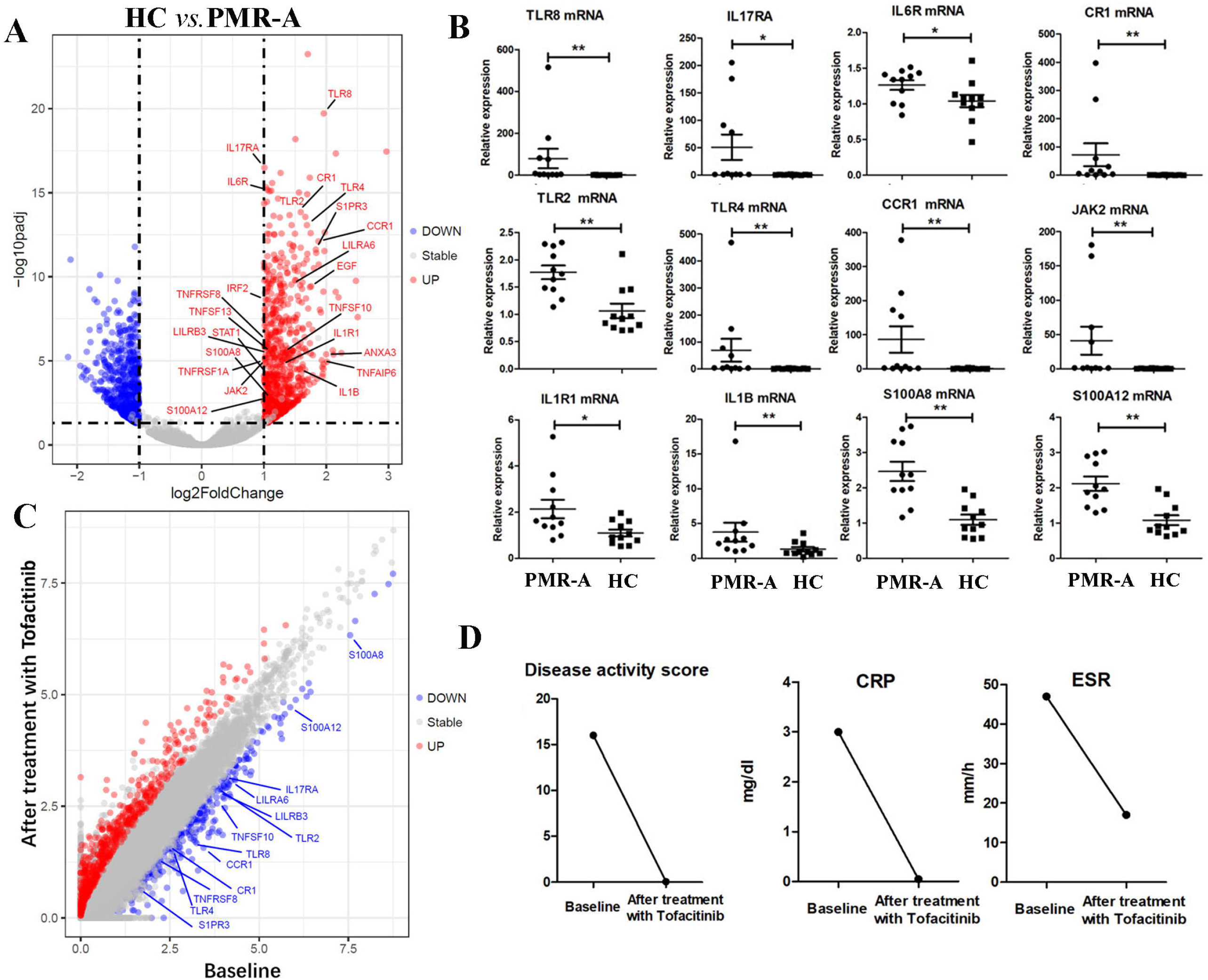
An abundant inflammatory genes which may activate JAK signaling. **(A)** Volcano results demonstrated the many genes expression were significantly increased, for example, TLR8, IL17RA, IL6R, CR1, TLR2, TLR4, CCR1,JAK2, IL1R1, IL1B, S100A8,and S100A12 (PMR-A, n=11 *vs*. HC n=20). **(B)** They were all increased in patients with PMR-A by Real-Time PCR analysis. Comparisons between two groups were performed using the nonparametric Mann-Whitney test (\**p*<0.05, \*\**p*<0.01). **(C)** Many genes were decreased in one representative patient with PMR. **(D)** She had a significant decrease in PMR disease score, CRP, and ESR after the treatment with tofacitinib.

We also confirmed that the above genes were all increased via Real-Time PCR (Fig. 2B). They can activate the JAK-signal transducer and activator of transcription (STAT) pathway[8-18]. They were also related to the function of monocytes, T cells, and neutrophils. The JAK-STAT pathway may be crucial in the pathogenesis of PMR.

### Tofacitinib therapy for patients with PMR

As we know, glucocorticoids are the first-line drugs in the treatment of PMR. However, they come with lots of side effects. Zhang et al. demonstrated that JAK1 and JAK3 were involved in chronic inflammation of media and large arteries in an animal model of giant cell arteritis (GCA), JAK inhibitor (JAKi) tofacitinib effectively suppressed innate and adaptive immunity in the vessel wall by reducing the proliferation rates of lesional T cells and their interferon-γ, IL-17, and IL-21 expressions[19]. We demonstrated that the expression of JAK2 was increased in PBMCs from patients with PMR (Figure 1B, C). Interestingly, Tofacitinib was tried in 3 patients with PMR as a steroid-sparing agent (https://isr.ie/abstracts/a-review-of-patients-started-on-janus-kinase-inhibitor-jaki-in-university-hospital-waterford/). Another JAKi Baricitinib lowered rapidly disease activity and exerted a significant steroid-sparing effect in 6 cases with refractory PMR and/or GCA [20]. JAKi appears as an appealing option for treating patients with PMR if they have refractory disease course or other diseases such as diabetes mellitus or osteoporosis which is relatively contraindicated for use of glucocorticoids. To some extent, RA and PMR are two inflammatory diseases with similar manifestations. JAKi was used for the treatment of RA. JAKi may be one choice for PMR. In our study, five patients with PMR refused to take steroids for fear of their side effects, because four patients have osteoporosis. They hopefully have a quick remission but no steroid or no injection such as tocilizumab. They agree with oral Tofacitinib therapy. Dramatically, their pain disappeared and many genes were decreased after tofacitinib therapy. One representative case is shown in Fig. 2C, D. There were no adverse events such as upper respiratory infection or herpes zoster during the 3–6-month period of treatment with tofacitinib.

Furthermore, the gene expression profiles of patients with PMR that were in remission after tofacitinib therapy (the PMR-C group) differed from those of patients in the PMR-A group and were similar to those of HC group (Fig. 1B). These results suggested that patients with PMR-C had attained complete serological remission after tofacitinib therapy.

## Discussion

PMR is a classic inflammatory disease with an acute onset but an unknown cause. IL-6 is one of the key pro-inflammatory cytokines involved in the pathogenesis of PMR. IL-6 and IL-1β were increased in the inflamed trapezius and lateral femoral muscles of PMR patients[8]. IL-6 can activate the JAK-STAT signaling via IL6R. Meanwhile, IL-1β binding to IL1R1 can form the high-affinity IL-1R complex, which mediates the Nuclear factor kappa B (NF-κB), Mitogen-activated protein kinase (MAPK), and JAK-STAT activation and promotes the inflammatory response[9]. TLR2, TLR4, and TLR8 were highly expressed in peripheral blood leukocytes, particularly in monocytes and macrophages. TLRs can lead to NF-κB activation, cytokine secretion, and the inflammatory response. TLR2/TLR4 in collaboration with the JAK-STAT signaling regulates endotoxemia and inflammation[10, 11]. STAT-1 was required for TLR8 transcriptional activity[12]. Furthermore, TLR4 was associated with increased susceptibility to PMR[21].

S100A8 and S100A12 are calcium-binding, zinc-binding proteins, and inflammatory mediators. S100A8 can induce the JAK1/2-dependent signaling[13]. S100A12 was induced by IL-6 via JAK signaling[14]. CCR1 and CR1 play critical roles in the process of inflammation. CCR1 was involved in the regulation of smoke-induced inflammation via JAK/STAT3 signaling[15]. CCR1 may also play a role in macrophage and endothelial cell infiltration in experimental arthritis models partly through activating JAK signaling[16]. Complement activation can be induced during inflammation, which is further normalized by a JAK1/2 inhibitor[17]. The number of Th17 cells was increased in the blood of PMR patients. CD161^+^CD4^+^T cells, which are precursors of Th17 cells, differentiated into Th17 cells through STAT-1/STAT-3 phosphorylation[18]. We found that IL6R, IL1B, IL1R1, TLR2, TLR4, TLR8, S100A8, S100A12, CCR1, CR1, and IL17RA expression was increased in PBMCs from patients with PMR. These genes may be partly involved in the pathogenesis of PMR via JAK-STAT signaling.

JAK1 and JAK3 were involved in chronic inflammation of media and large arteries in an animal model of GCA, tofacitinib effectively suppressed innate and adaptive immunity in the vessel wall [19]. Furthermore, five patients with PMR had a good response to tofacitinib therapy in our study. We have registered a clinical trial titled “a prospective study on the efficacy and safety of tofacitinib in the treatment of PMR” (ChiCTR2000038253) on the website (http://www.chictr.org.cn/). More data will be published in the future.

Our study has some limitations. The number of patients was small. We do not finish the functional experiment of candidate genes involved in the pathogenesis of PMR. PMR is closely associated with GCA. The patients with PMR in this study did not simultaneously have a GCA. Further studies are needed to investigate the clinical role and mechanism of action of candidate genes in PMR.

## Conclusion

Many inflammatory genes associated with JAK signaling are highly enriched in PBMCs from patients with PMR. Tofacitinib, a pan JAK inhibitor effectively treated the PMR patients with clinical remission and sharply decreased in CRP and ESR in this pre-clinical trial study. Tofacitinib may effectively treat PMR patients.

## Data Availability

All data produced in the present study are available upon reasonable request to the authors.

## Data availability statement

All the data can be available upon reasonable request.

